# Novel Autosegmentation Spatial Similarity Metrics Capture the Time Required to Correct Segmentations Better than Traditional Metrics in a Thoracic Cavity Segmentation Workflow

**DOI:** 10.1101/2020.05.14.20102103

**Authors:** Kendall J. Kiser, Arko Barman, Sonja Stieb, Clifton D. Fuller, Luca Giancardo

## Abstract

**Introduction:** Automated segmentation templates can save clinicians time compared to de novo segmentation but may still take substantial time to review and correct. It has not been thoroughly investigated which automated segmentation-corrected segmentation similarity metrics best predict clinician correction time.

**Materials and Methods:** Bilateral thoracic cavity volumes in 329 CT scans were segmented by a UNet-inspired deep learning segmentation tool and subsequently corrected by a fourth-year medical student. Eight spatial similarity metrics were calculated between the automated and corrected segmentations and associated with correction times using Spearman’s rank correlation coefficients. Nine clinical variables were also associated with metrics and correction times using Spearman’s rank correlation coefficients or Mann-Whitney U tests.

**Results:** The added path length, false negative path length, and surface Dice similarity coefficient correlated better with correction time than traditional metrics, including the popular volumetric Dice similarity coefficient (respectively ρ = 0.69, ρ = 0.65, ρ = –0.48 versus ρ = –0.25; correlation p values < 0.001). Clinical variables poorly represented in the autosegmentation tool’s training data were often associated with decreased accuracy but not necessarily with prolonged correction time.

**Discussion:** Metrics used to develop and evaluate autosegmentation tools should correlate with clinical time saved. To our knowledge, this is only the second investigation of which metrics correlate with time saved. Validation of our findings is indicated in other anatomic sites and clinical workflows.

**Conclusion:** Novel spatial similarity metrics may be preferable to traditional metrics for developing and evaluating autosegmentation tools that are intended to save clinicians time.

## INTRODUCTION

The advent of deep learning-based segmentation algorithms is expanding the range of automated segmentation (autosegmentation) use to clinical tasks and research questions that demand previously unattainable accuracy or reliability. Autosegmentation algorithms may soon assist neurologists to localize ischemic cores during a code stroke^1,2^ or anticipate Parkinson’s disease onset in an outpatient setting.^3^ They may inform 3-D printed implant designs for orthopedists^4,5^ or highlight posterior segment lesions^6–8^ for ophthalmologists. They may help neurosurgeons spare microvessels,^9^ outline catheters for radiation oncologists during MRI-guided brachytherapy,^10^ or characterize vocal fold mobility for otorhinolaryngologists.^11^ Dedicated imaging specialists – radiologists and pathologists – are likely to identify even more autosegmentation uses than clinicians whose primary clinical domain is not imaging. For example, segmenting regions-of-interest is a necessary step prior to extraction of quantitative imaging biomarkers (“radiomics” features) known to harbor information respecting disease prognoses and treatment response probabilities.^12^ Radiomics feature computation methods were recently standardized,^13^ overcoming a significant obstacle to clinical implementation. In the future, reviewing and vetting autosegmented regions-of-interest prior to radiomics analyses could become part of routine radiology.^14^

Autosegmentations are useful if they obviate the need for a clinician to delineate segmentations de novo, which can be time-consuming^4,15–18^ and inconsistent^19–23^ between observers and within the same observer at different time points. Several studies confirm that clinicians can save time from leveraging autosegmentation templates compared to de novo segmentation,^15,24–29^ but in many circumstances the time required for clinicians to review and correct autosegmentations is still substantial. For example, during online adaptive/stereotactic MRI-guided radiotherapy,^30^ radiation oncologists must carefully correct cancer and normal anatomy autosegmentations while a patient waits immobilized in the treatment device. Cardiologists may spend just as long to review and correct cardiac ventricle autosegmentations as segmenting them de novo.^18^ Plastic surgeons can implant facial trauma repair plates faster with autosegmentation-based 3D-printed mandibular templates than without them,^31^ but autosegmentation review still consumes time in an urgent setting. Whenever autosegmentation algorithms are deployed to save clinical time, the metrics used to assess them should capture an expected time-savings benefit. Algorithm development should be optimized and evaluated by whatever metric or metrics best predict time savings.

Autosegmentations are usually compared with a reference segmentation by spatial similarity metrics that compare 1) volumetric overlap between an autosegmented structure and the same manually segmented structure,^4,6,10,11,16,22-26,28,29,32–47^ such as the volumetric Dice similiarity coefficient/index^48^ (DSC), or 2) geometric distance between two structures’ surfaces,^4,10,22,24–26,28,32-35,37,40,42,43,46,47^ such as the Hausdorff distance^49^ (HD), or 3) structure centricity,^32,39^ such as differences between centers-of-mass. Critically, these metrics do not necessarily correlate with time savings in clinical practice.^40,50^ To our knowledge, the only investigation of *which* metrics best predict the time clinicians spend correcting autosegmentations was published in 2020 by Vaassen et al.^28^

Vaassen et al. compared automatically generated and manually corrected thoracic structure segmentations in 20 CT cases acquired from patients with non-small-cell lung cancer (NSCLC). They found that the “added path length” (APL; a novel metric they introduced) and the surface Dice Similarity Coefficient (a novel metric introduced by Nikolov et al.^51^) correlated better with the time it took a clinician to review and correct autosegmentations than other metrics that are popular for autosegmentation evaluation. Here, we corroborate and extend their findings. We also experiment with APL by calculating variations of it, which we term the false negative path length (FNPL) and false negative volume (FNV). We correlate the APL, FNPL, FNV, surface DSC, volumetric DSC, Jaccard index (JI), average surface distance (ASD), and HD metrics calculated between automatically generated and manually corrected thoracic cavity segmentations with time required for correction. We contribute evidence that the surface DSC may be superior to popular volumetric DSC for optimizing autosegmentation algorithms. We also investigate how anatomic and pathologic variables impact autosegmentation correction time. In the process we have generated a library of 402 expert-vetted left and right thoracic cavity segmentations, as well as 78 pleural effusion segmentations, which we made publicly available through The Cancer Imaging Archive^52^ (TCIA) at doi:10.7937/tcia.2020.6c7y-gq39.^53^ The CT scans on which the segmentations were delineated are likewise publicly available^54^ from TCIA.

## MATERIALS AND METHODS

### CT Datasets

A collection of four hundred twenty-two CT datasets acquired in Digital Imaging and Communications in Medicine (DICOM) format from patients with NSCLC was downloaded from NSCLC-Radiomics,^54^ a TCIA data collection, in January 2019. Accompanying clinical data in tabular format and gross tumor volume segmentations available for a subset of cases were also downloaded. CT scans were converted from DICOM to Neuroimaging Informatics Technology Initiative (NIfTI) format using a free program called “dcm2niix.”^55,56^ Four-hundred-two CT datasets were successfully converted and subsequently underwent autosegmentation and manual correction.

### Segmentations

We leveraged a publicly available, UNet-inspired deep learning autosegmentation algorithm^57^ to segment lungs in the 402 CT datasets described above. This algorithm was trained to segment bilateral lungs (under a single label) with approximately 200 CTs acquired in patients who – importantly – did not have lung cancer. A fourth-year medical student reviewed and corrected the autosegmentations using an image segmentation software called ITK-SNAP v 3.6.^58^ The corrections included the bilateral thoracic cavity spaces that healthy lung parenchyma normally occupies, but in our dataset were occasionally occupied by atelectatic parenchyma, tumor, pleural effusion, or other changes. Because the idea to capture correction time and correlate it with autosegmentation similarity metrics developed after this project had commenced, the medical student recorded the time it took to correct autosegmentations for only 329 of 402 corrected cases. Specifically, correction times comprised the times required to load autosegmentations, correct them slice-wise with size-adjustable brush and interpolation tools, and save the corrections. Because the autosegmentation algorithm was trained on scans without cancer but deployed on scans with NSCLC, its accuracy varied with the severity of disease-induced anatomic change in each case. For example, cases with massive tumors or pleural effusions were sometimes poorly autosegmented, whereas cases with minimal anatomic changes were autosegmented well. This effectively simulated a range of major and minor manual corrections. Subsequently, the medical student’s manually corrected segmentations were vetted and corrected as necessary by a radiation oncologist or a radiologist. The 402 physician-corrected thoracic cavity segmentations – so named to reflect inclusion of primary tumor and pleural pathologies in the thoracic cavity rather than lung parenchyma alone – have been made publicly available at TCIA.^53^

### Metrics

Automated and corrected segmentations were compared by the volumetric DSC, the JI, the surface DSC at 0mm, 4mm, 8mm, and 10mm tolerances, the APL, the FNPL, the FNV, the 100^th^, 99^th^, 98^th^ and 95^th^ percentile HDs, and the ASD. Each metric is illustrated in Figure 1. The volumetric DSC is twice the overlap between volumes A and B, divided by their sum. A DSC of 1 indicates perfect overlap while 0 indicates no overlap. The JI is a related volumetric measure and is the overlap between volumes A and B divided by their union. The DSC and JI converge at 1.^59^ The surface DSC is calculated by the same formula as the volumetric DSC, but its inputs A and B are the segmentations’ surface areas rather than their volumes. To permit small differences between surfaces to go unpunished, Nikolov et. al programmed a tolerance parameter: if points in two surfaces are separated by a distance that is within the tolerance parameter, they are considered part of the intersection of A and B. The APL is the number of pixels in the corrected segmentation surface (edge) that are not in the autosegmentation surface^28^. We experiment with metrics related to the APL that we term the FNPL and the FNV. The FNPL is the APL less the pixels from any edits that shrink the autosegmentation. That is, edits that erase pixels from the autosegmentation volume are excluded. The FNV is the number of pixels in the corrected segmentation volume that are not in the autosegmentation volume. Python code we developed to calculate the APL, FNPL, and FNV has been made available at GitHub at https://github.com/kkiser1/Autosegmentation-Spatial-Similarity-Metrics. The Hausdorff distance calculates the minimum distance from every point in surface A to every point in surface B, and vice versa, arranges all distances in ascending order, and returns the maximum distance (100^th^ percentile) or another percentile if so specified (e.g. 95^th^ percentile). The ASD calculates the average of the minimum distances from every point in surface A to every point in surface B, and vice versa, and returns the average of the two average distances. All metric calculations were made using custom Python scripts that leveraged common scientific libraries.^51,60–62^

**Figure 1:**
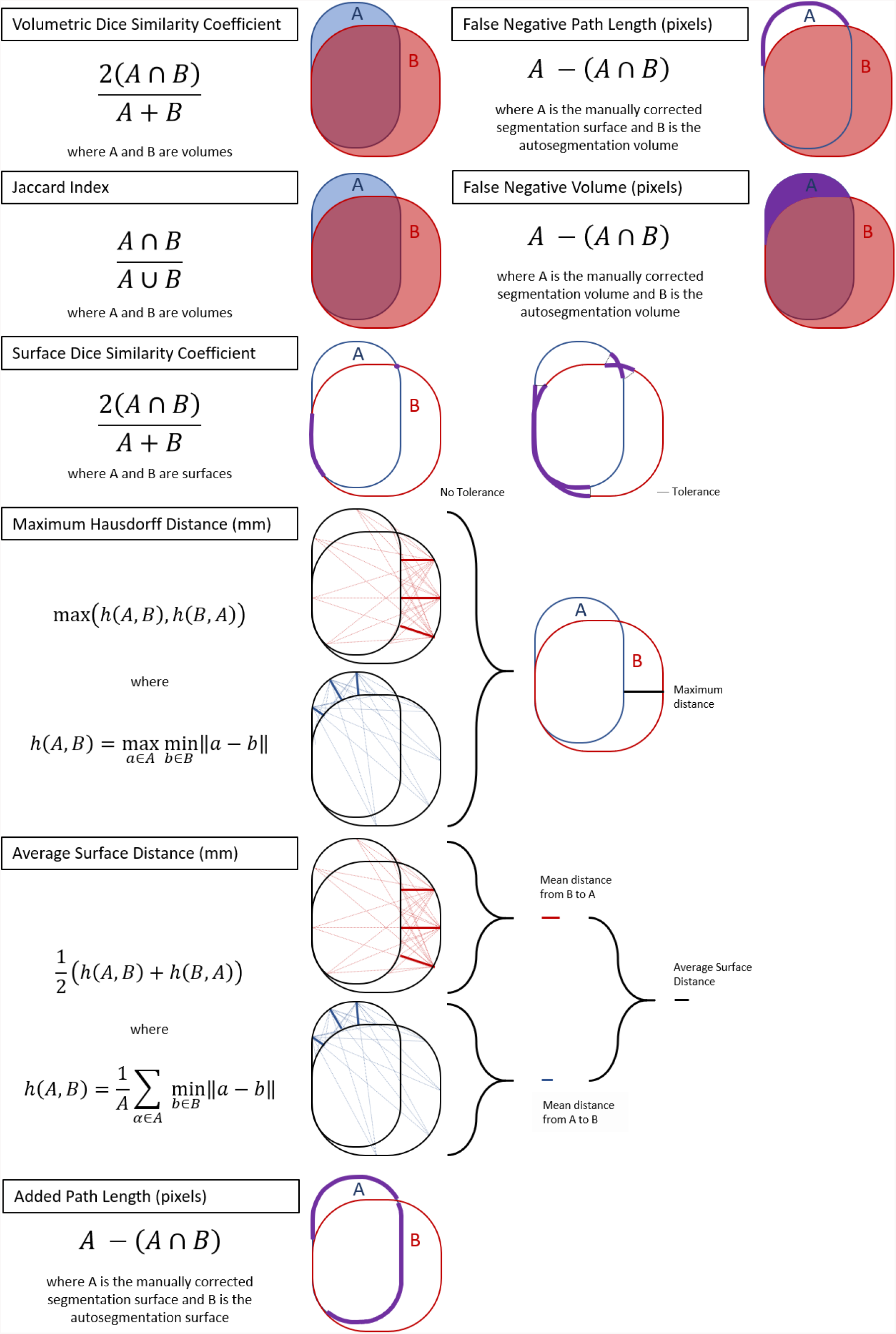
Eight metrics for evaluating spatial similarity between segmentations. Traditional (volumetric DSC, Jaccard index, Hausdorff distance, and average surface distance) or novel (surface DSC, added path length, false negative path length, false negative volume), were used to compare autosegmentations with manually corrected segmentations. The surface DSC calculation permits a tolerance parameter whereby non-intersecting segments of surfaces A and B that are separated by no more than the parameter distance are considered part of the intersection between A and B. The Hausdorff distance illustration and equation represent the 100^th^ percentile (maximum) distance but can be adapted to any other percentile distance.

### Clinical variables

To describe clinical variation in the NSCLC-Radiomics CT datasets and study the effects of variation in tumor volume, tumor laterality and location, pleural effusion presence, pleural effusion volume, and thoracic cavity volume on autosegmentation spatial similarity metrics and on manual correction time, we collected these variables for each case. Furthermore, we studied how primary tumor stage, tumor overall stage, and tumor histology associated with accuracy and correction time, but these variables were already collected in the NSCLC-Radiomics collection^54^ in a spreadsheet named “NSCLC Radiomics Lung1.clinical-version3-Oct 2019.csv.” Left and right thoracic cavity volumes were collected from physician-vetted thoracic cavity segmentations using ITK-SNAP. Tumor volume and laterality were collected by referencing primary gross tumor volume segmentations (“GTV-1”) and other tumor volume segmentations available from the NSCLC-Radiomics data collection.^54^ Tumor location was classified as central, peripheral, or pan. There is not consensus in radiotherapy literature regarding the definition of centrality^63^; we used a definition based off that provided by the International Association for the Study of Lung Cancer^64^: tumors located within 2 cm of the proximal bronchial tree, spinal cord, heart, great vessels, esophagus, or phrenic nerves and recurrent laryngeal nerves and spanning up to 4 cm from these structures were classified as central. Tumors that were not within 2 cm of any central structure were classified as peripheral. Tumors within the central territory that extended further than 4 cm from central structures were classified as pan. The presence or absence of pleural effusion in each subject was noted by a medical student, and effusions were contoured by the student. Pleural effusion segmentations were reviewed and corrected by a radiologist. Pleural effusion volumes were collected from physician-vetted segmentations using ITK-SNAP.

### Statistics

We correlated eight autosegmentation spatial similarity metrics with the time expended to correct the autosegmentations. Segmentation correction time, volumetric DSC, surface DSC, JI, APL, FNPL, FNV, HD, and ASD distributions were assessed for normality using the Shapiro-Wilks test.^65^ The null hypotheses (that distributions were normal) were rejected in each case (p values < 0.001). Additionally, we sought to describe the influence of common disease-induced anatomic changes on the accuracy of the UNet autosegmentation algorithm. The null hypotheses that tumor, thoracic cavity, and pleural effusion volumes distributions were normal were likewise each rejected (p values < 0.001). Therefore, non-parametric statistical tests were employed. Pairwise Spearman’s rank correlation coefficients^66^ described linear associations between spatial similarity metrics and correction times. The pairwise Mann-Whitney U test^67^ assessed significant differences between numeric variable distributions stratified by a categorical variable with two categories, and was preceded by the Kruskal-Wallis^68^ test if the categorical variable had three or more categories and the Kruskal-Wallis result was significant. A significance threshold of α = 0.05 was used, and Bonferroni corrections^69^ were assessed to account for multiple comparisons as needed. Statistics were computed in Python using the SciPy library.^60^

## RESULTS

Four-hundred and two thoracic cavity segmentations were automatically generated and corrected manually (Figure 2). Correction times were recorded in 329 cases. Among these cases, median right and left corrected thoracic cavity volumes were 2220 cm^3^ and 1920 cm^3^, respectively (Figure 3a). Tumor overall stage was I in 10% of cases (33/329), II in 21% of cases (69/329), IIIA in 27% of cases (88/329), and IIIB in 42% of cases (139/329). Tumor stage was T1 in 24% of cases (78/329), T2 in 34% of cases (112/329), T3 in 13% of cases (42/329), and T4 in 29% of cases (97/329). Primary lung tumors were in the right hemithorax in 58% of cases (191/329) and in the left hemithorax in 42% of cases (138/329). Tumor locations were classified as central in 24% of cases (80/329), peripheral in 33% of cases (108/329), and pan in 43% of cases (141/329). Median tumor volumes were 29 cm^3^, 17 cm^3^, 2 cm^3^, 4 cm^3^, 3 cm^3^, and 6 cm^3^ for GTV1 through GTV6, respectively (Figure 3b). Among 298 cases with recorded autosegmentation time and available tumor histology, the histology was squamous cell carcinoma in 40% of cases (120/298), large cell carcinoma in 30% of cases (90/298), adenocarcinoma in 14% of cases (43/298), and not otherwise specified in 15% of cases (45/298). Among 59 cases with recorded autosegmentation correction times and a pleural effusion in at least one hemithorax, median right and left pleural effusion volumes were 53 cm^3^ and 51 cm^3^, respectively (Figure 3c).

**Figure 2:**
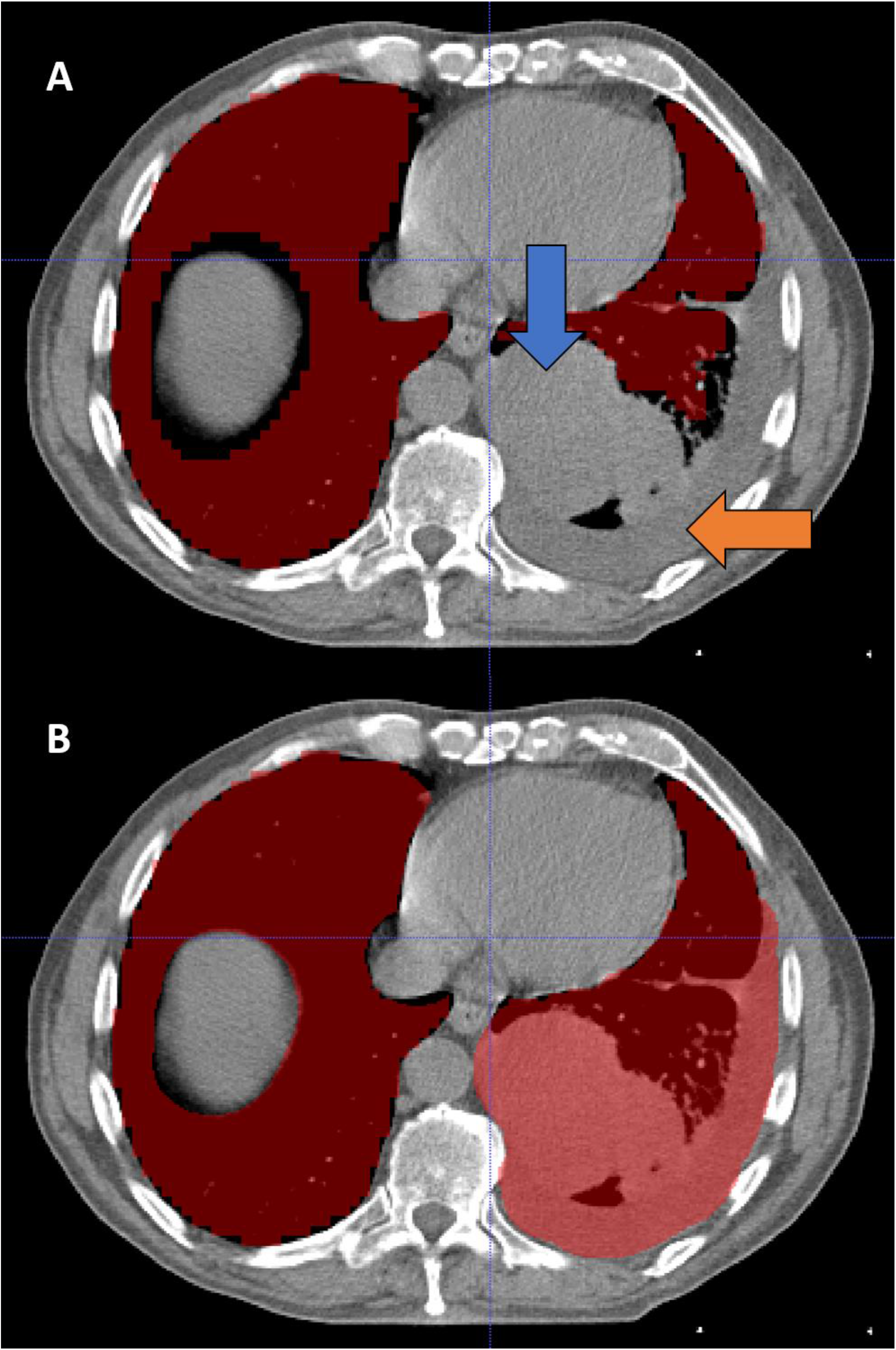
A) A deep learning algorithm segmented bilateral thoracic cavity volumes. Accuracy varied in the presence of disease-induced anatomic changes, exemplified by pleural effusion (orange arrow) and primary tumor (blue arrow). B) A fourth-year medical student corrected the autosegmentations.

**Figure 3:**
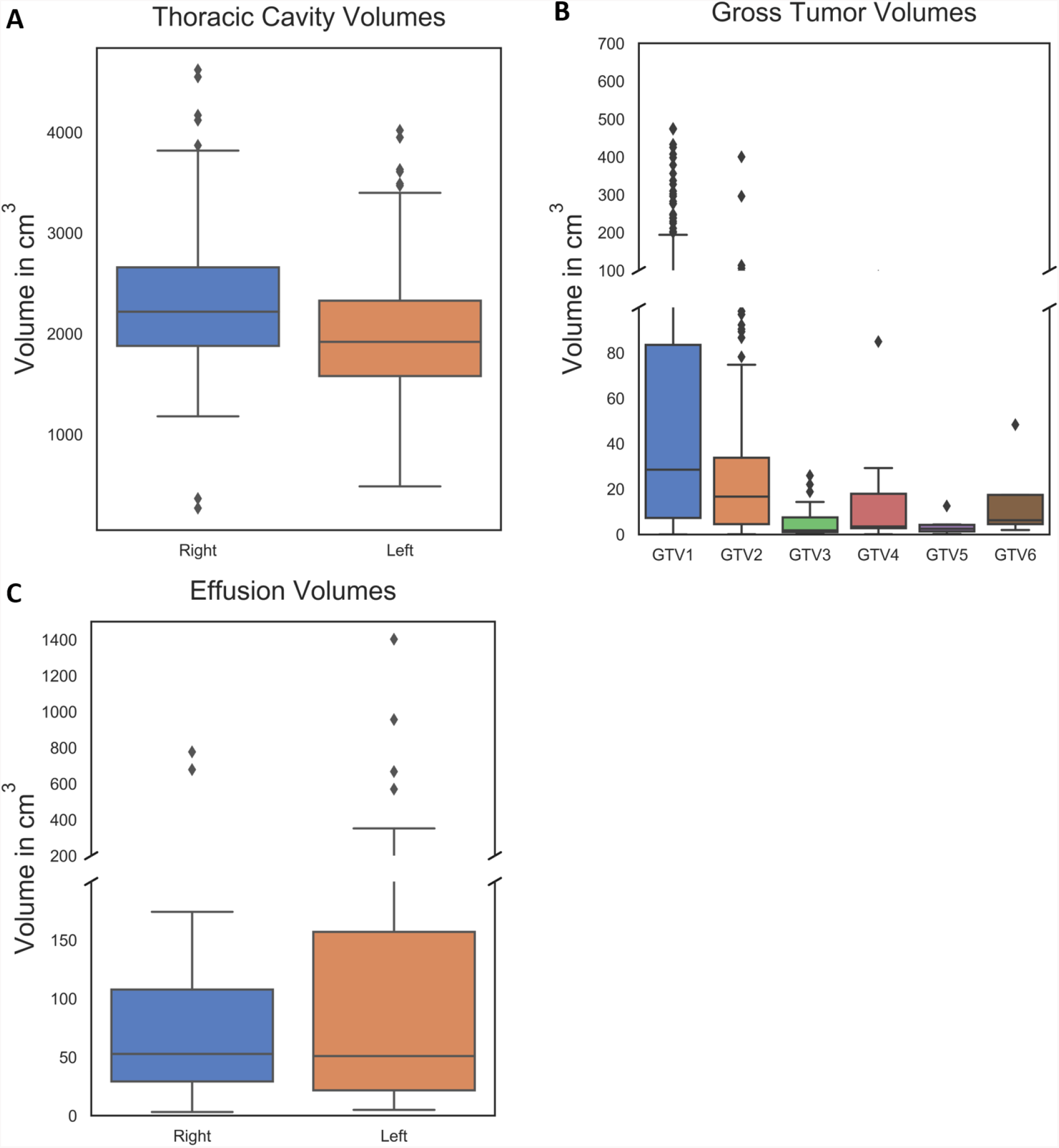
A) Right and left thoracic cavity volumes in cases with a recorded autosegmentation correction time (n = 329). Volumes were collected after autosegmentation correction by a medical student and subsequent vetting by a physician. B) Gross tumor volumes as delineated in “RTSTRUCT” segmentation files available from The Cancer Imaging Archive NSCLC-Radiomics data collection.^54^ “GTV1” denotes the primary tumor volumes (n = 328), whereas “GTV2” through “GTV6” denote secondary tumor volumes that were occasionally present. Usually, the latter were clusters of mediastinal nodes. Because the mediastinum is not part of the lung nor the space healthy lung usually occupies, correlations with tumor volume consider only “GTV1,” not the sum of “GTV1” through “GTV6.” C) Right and left pleural effusion volumes in cases with a pleural effusion and a recorded thoracic cavity autosegmentation correction time (n = 59). These were delineated de novo by a medical student (rather than corrected from an autosegmentation template) and subsequently vetted by a radiologist.

Anatomic changes caused by disease significantly influenced the autosegmentation algorithm’s similarity to manually corrected segmentations, but worse similarity did not always result in longer correction times. Tumor location (central, peripheral, or pan) was associated with similarity by several metrics (e.g. volumetric DSC for central tumors: 0.963, pan tumors: 0.945; p < 0.001), but there were no significant differences in correction time between central (median 18.61 min), peripheral (median 19.01 min), or pan (median 18.83 min) tumors (p = 0.24). Primary tumor volume correlated moderately with several similarity metrics but not with correction time (p = 0.15). Like tumor location, the presence of pleural effusion was associated with significantly worse similarity by all metrics (p values < 0.001), but this did not significantly prolong manual correction times (median with effusion: 19.13 min, without effusion: 18.71 min; p = 0.18). Furthermore, pleural effusion volume correlated weakly with volume overlap metrics (i.e. volumetric DSC, JI) but not with correction time (p = 0.20). Neither metric nor correction time distributions were significantly different between tumor histologies.

Few clinical variables were significantly associated with correction time. Autosegmentations delineated on CT scans with T4 tumors took marginally but significantly longer to correct (median 20.82 min) than those on CTs with T1 (median 19.0 min), T2 (median 18.13 min), or T3 (median 18.30 min) tumors (p values ≤ 0.01). Interestingly, the only metrics that captured significant differences between cases with T4 tumors and cases with any other T stage tumor were the maximum HD (median T4: 56 mm, median T3: 70 mm; p = 0.02), FNV (median T4: 104,991 pixels, median T1: 89,334 pixels; p = 0.001), FNPL (median T4: 61,928 pixels, median T2: 56,612 pixels, median T1: 57,489 pixels; p values ≤ 0.006), and APL (median T4: 69,707 pixels, median T2: 61,553 pixels, median T1: 61,788 pixels; p values ≤ 0.002). Autosegmentations on CT scans with overall stage II tumors took significantly less time to correct (median 13.53 min) than those from CTs with stage I (median 19.08 min), stage IIIA (median 18.94 min), or stage IIIB tumors (median 19.83 min) (p values ≤ 0.04). Only the surface DSC at 0 mm tolerance, FNPL, and APL captured a significant difference between these groups (surface DSC median for stage II: 0.77 vs. stage I: 0.70, stage IIIA: 0.70, stage IIIB: 0.70; p values ≤ 0.004; FNPL median for stage II: 48,262 pixels vs. stage I: 62,698 pixels, stage IIIA: 58,211 pixels, stage IIIB: 58,863 pixels; p values ≤ 0.004, APL median for stage II: 52,260 pixels vs. stage I: 67,446 pixels, stage IIIA: 62,986 pixels, stage IIIB: 67,054 pixels; p values ≤ 0.001). Of the quantitative clinical variables, total thoracic cavity volume was the only significant correlate with correction time (ρ = 0.19, p < 0.001).

Linear correlations between autosegmentation spatial similarity metrics and correction times were also evaluated. Correction time and metric distribution summary statistics are reported in Table 1. All metrics had statistically significant correlations with correction time (p values < 0.05), but the strength of these correlations varied from strongest to weakest as follows: APL (ρ = 0.69, p < 0.001), FNPL (ρ = 0.65, p < 0.001), surface DSC at 0mm tolerance (ρ = –0.48, p < 0.001), FNV (ρ = 0.40, p < 0.001), JI (ρ = –0.26, p < 0.001), volumetric DSC (ρ = –0.25, p < 0.001), ASD (ρ = 0.24, p < 0.001), surface DSC at 4mm tolerance (ρ = –0.23, p < 0.001), 95^th^ percentile HD (ρ = 0.20, p < 0.001), surface DSC at 8mm tolerance (ρ = –0.20, p < 0.001), surface DSC at 10mm tolerance (ρ = –0.19, p < 0.001), 98^th^ percentile HD (ρ = 0.17, p = 0.002), 99^th^ percentile HD (ρ = 0.11, p = 0.04), and maximum HD (ρ = 0.11, p = 0.05). Correction time correlations with conformality metrics (volumetric DSC, JI, and best-performing surface DSC) are visualized in Figure 4, with surface distance metrics (best-performing HD and ASD) in Figure 5, and with pixel count metrics (APL, FNPL, FNV) in Figure 6.

**Figure 4:**
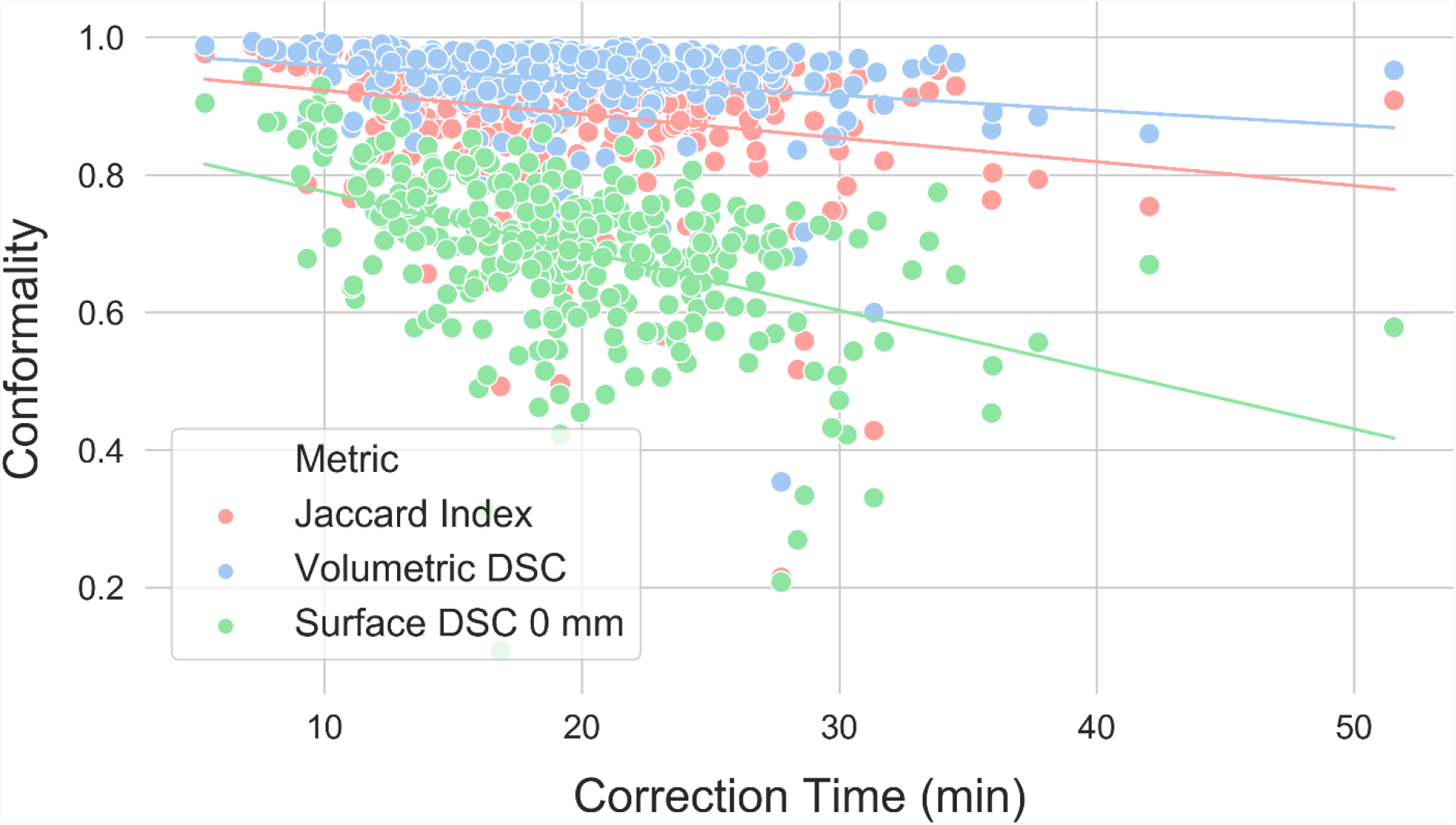
Correlations between correction time and conformality metrics. The surface Dice similarity coefficient at 0mm tolerance correlated more strongly with correction time (ρ = –0.48, p < 0.001) than any other conformality, surface distance, or pixel metric except the added path length and false negative path length. Other conformality metrics correlated poorly (Jaccard index: ρ = –0.26, p < 0.001; volumetric Dice similarity coefficient: ρ = –0.25, p < 0.001).

**Figure 5:**
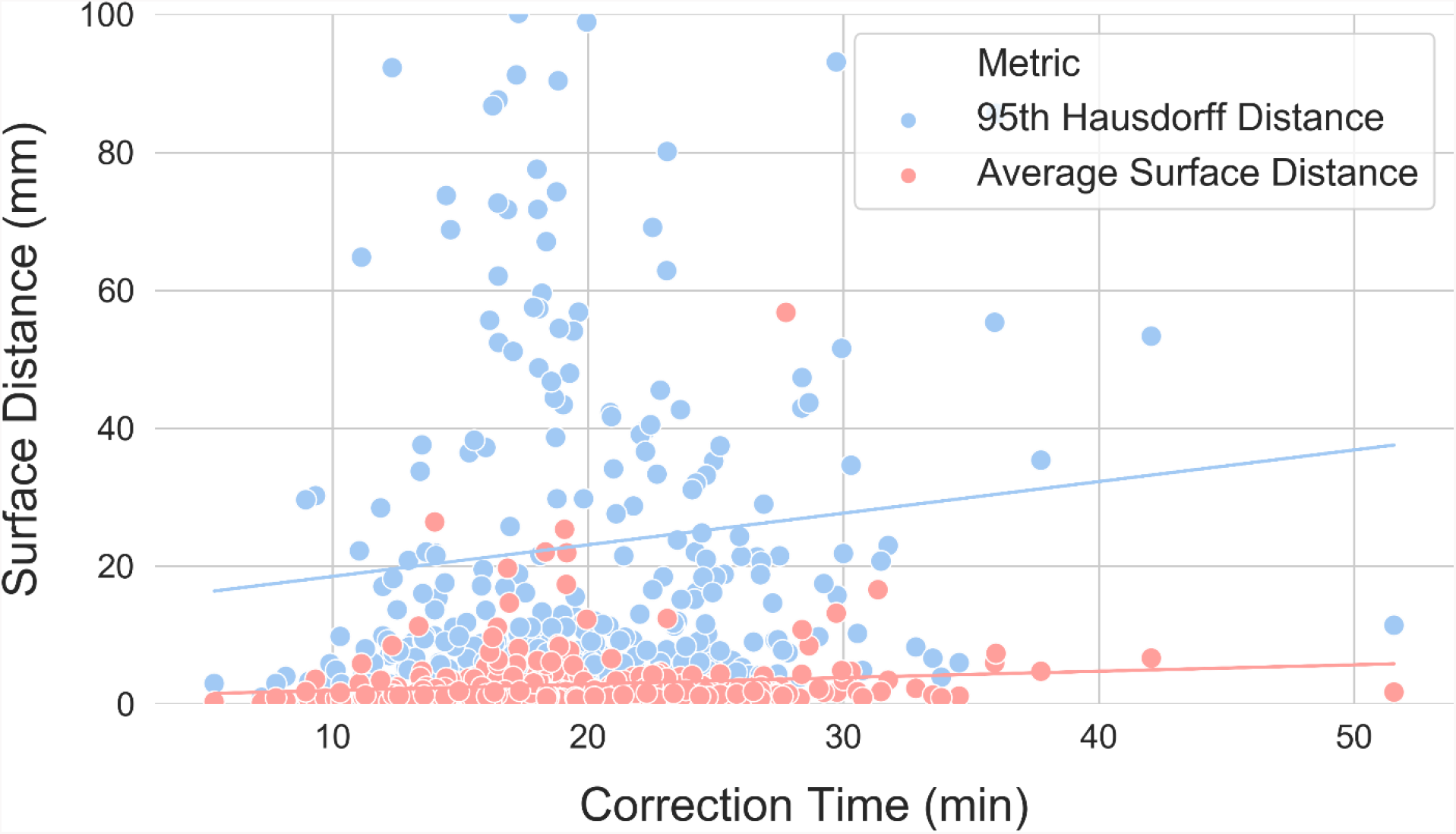
Correlations between correction time and surface distance metrics. For visual clarity, only the average surface distance (ρ = 0.24, p < 0.001) and the 95th percentile Hausdorff distance (ρ = 0.20, p < 0.001) are displayed, which are the two best-performing surface distance metrics. The y axis maximum has been limited to better visualize the distributions, excluding ten 95^th^ percentile Hausdorff distance points that exceeded 100 mm. As a class, surface distance metrics were poorer correlates with correction time than conformality or pixel metrics.

**Figure 6:**
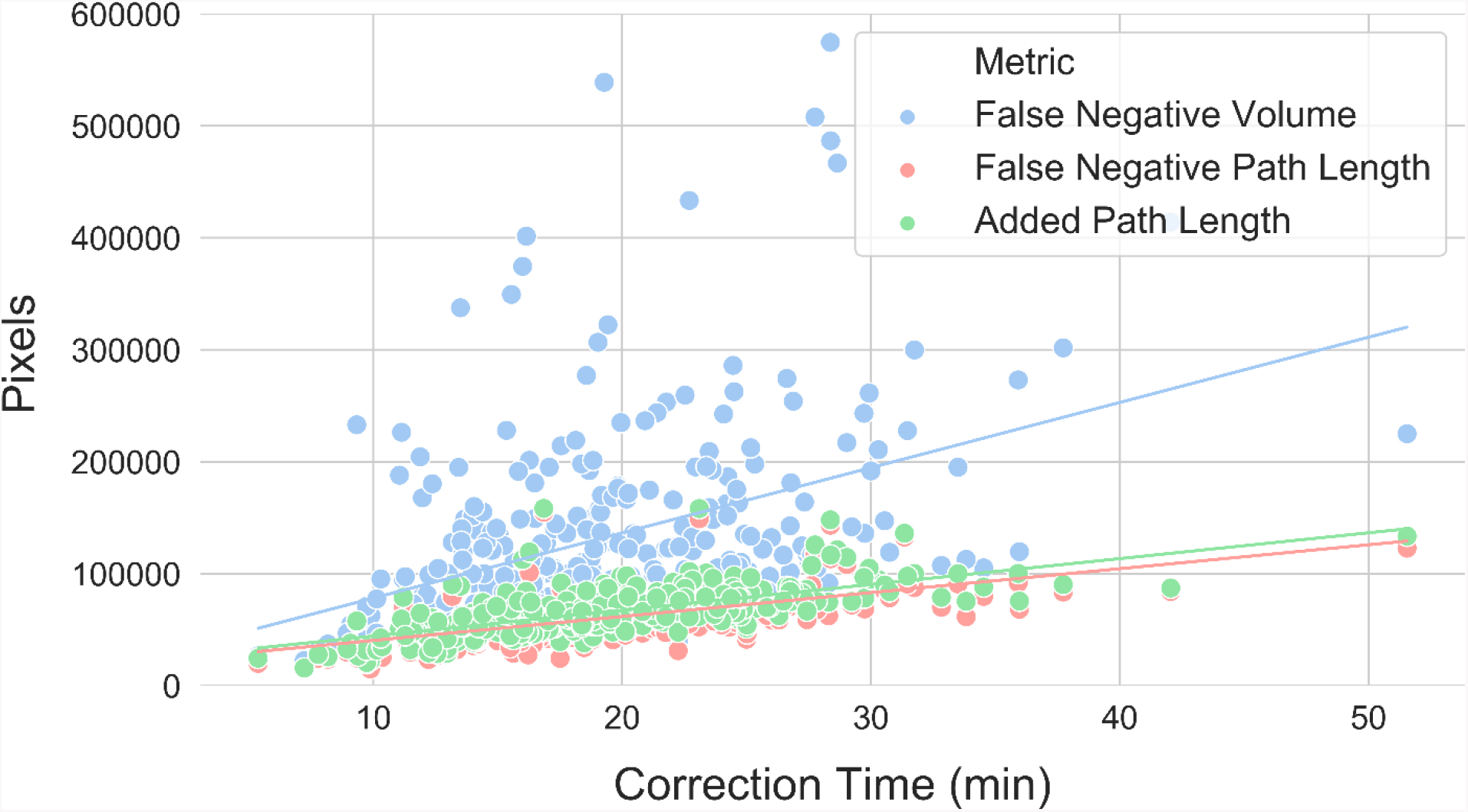
Correlations between correction time and pixel count metrics. The added path length correlated better with correction time than any other metric (ρ = 0.69, p < 0.001), while the false negative path length (ρ = 0.65, p < 0.001) and false negative volume (ρ = 0.40, p < 0.001) were respectively the second and fourth-best performing metrics. The y axis maximum has been limited to better visualize the distributions, excluding three false negative volume points between 600,000 and 1,000,000 pixels.

**Table 1:**
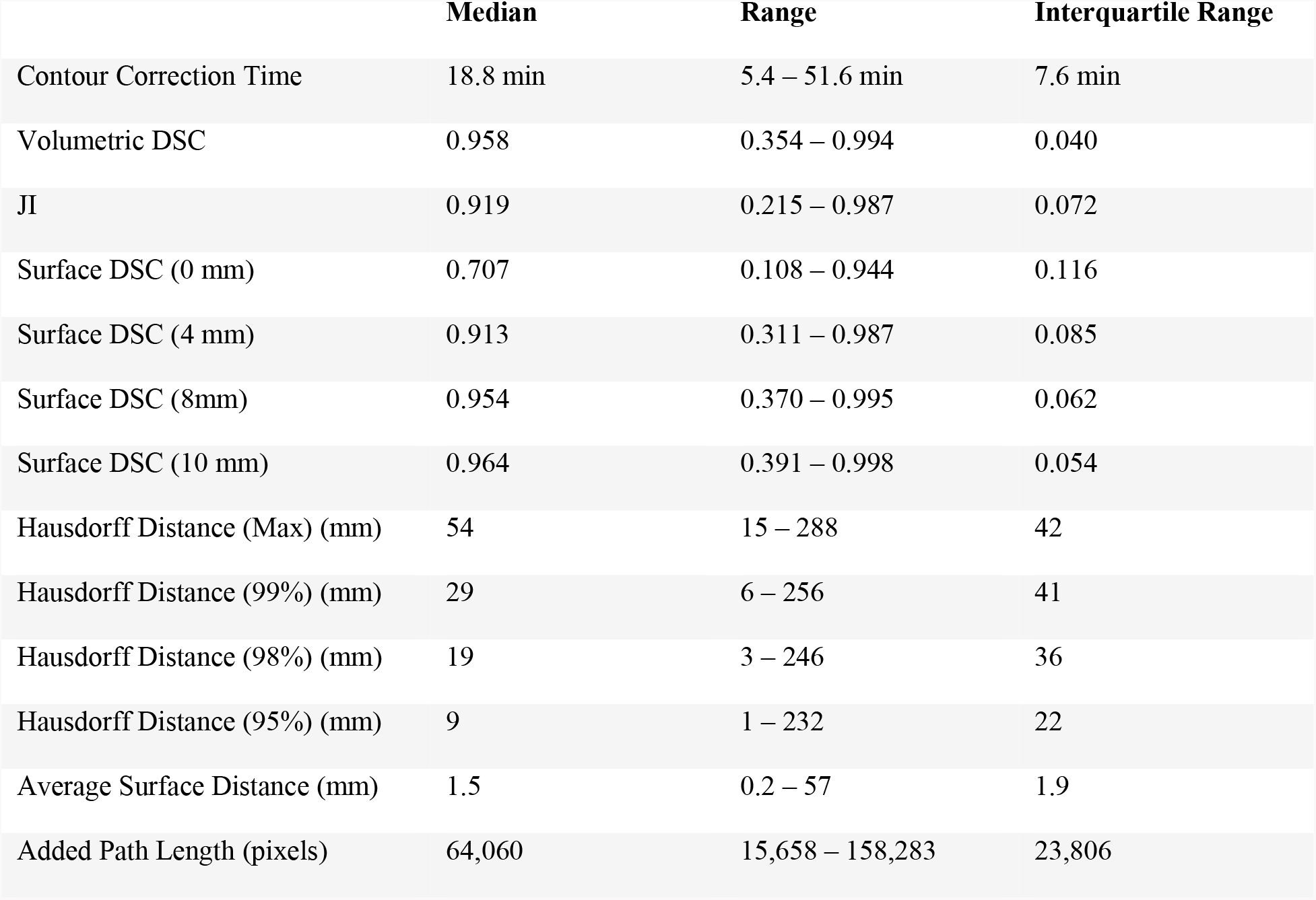
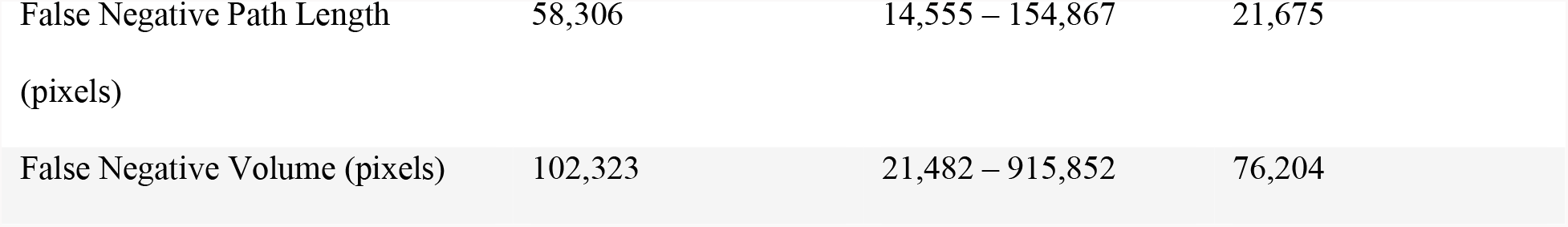
Eight spatial similarity metrics were calculated between autosegmented and manually corrected bilateral thoracic cavity segmentations (n = 329). Two metrics – the surface Dice similarity coefficient and the Hausdorff distance – were calculated with different parameters. Table values are the median, range, and interquartile range for each of these distributions.

Secondary regression analyses were performed between autosegmentation spatial similarity metrics and correction times after stratifying by clinical variables known to have significant relationships with correction times (i.e. T stage, overall stage, and total thoracic cavity volume; thoracic cavity volume was transformed to a categorical variable by binning volumes by quartile). The APL, FNPL, and surface DSC at 0 mm remained highly significant correlates with correction time in every T stage, overall stage, and thoracic volume quartile subgroup (p values < 0.001) except the stage IIIA subgroup, in which only the APL and FNPL (but not the surface DSC) were significant correlates. APL ρ correlation coefficients ranged from 0.60 – 0.80 and were the highest of all metrics in every subgroup except the thoracic cavity volume 1^st^ quartile subgroup, in which the surface DSC at 0 mm correlation coefficient was slightly stronger (ρ = –0.62).

## DISCUSSION

Autosegmentation algorithms can assist physicians in an increasing number of clinical tasks, but algorithms are evaluated by spatial similarity metrics that do not necessarily correlate with clinical time savings. The question of which metrics correlate best with time savings has not been thoroughly investigated. To our knowledge, ours is only the second and largest study described for this purpose. In thoracic cavity segmentations delineated on 329 CT datasets, we evaluated correlations between the time required to review and correct autosegmentations and eight spatial similarity metrics. We find the APL, FNPL, and surface DSC to be better correlates with correction times than traditional metrics, including the ubiquitous^4,6,10,11,16,22–26,28,29,32–47^ volumetric DSC. We find that clinical variables that worsen autosegmentation similarity to manually-corrected references do not necessarily prolong the time it takes to correct the autosegmentations. We also show that APL, FNPL, and surface DSC remain strong correlates with correction time even after controlling for clinical variables that *do* prolong correction time. Using the APL or surface DSC to optimize algorithm training – such as to compute a loss function^70,71^ – may make the algorithms’ outputs faster to correct. Using them to assess autosegmentation performance may communicate a more accurate expectation of the time needed to correct the autosegmentations. Notably, for any comparison of two segmentations where neither can be considered the reference standard, the surface DSC should be preferred to the APL. The surface DSC is directionless, but calculating the APL requires designating one segmentation as a standard. Autosegmentations that are optimized to save clinicians time may facilitate faster urgent and emergent interventions.^1,2^ They may decrease intraoperative overhead costs.^31^ They may be especially beneficial for treatment paradigms that demand daily image segmentation. For example, in an online adaptive MRI-guided radiotherapy workflow, autosegmentations for various anatomic structures are generated every day. Segmentation review occurs while the patient remains in full-body immobilization^30,72^. This creates a need for a metric to generate a “go/no-go” decision for real-time manual segmentation.^73^ Computing the APL between autosegmentations-of-the-day and the physician-approved segmentations from the previous day could signal to the radiation oncologist whether re-segmentation is likely feasible within the time constraints of online fractionation, or whether offline corrections are needed given patient time-in-device. Furthermore, optimized autosegmentation algorithms are foundational to unlocking the benefits of artificial intelligence in radiology; indeed, the Radiological Society of North America, National Institutes of Health, and American College of Radiology identify improved autosegmentation algorithms among their research priorities.^74^ These benefits include clinical implementation of radiomics-based clinical decision support systems. While not the only obstacle preventing implementing of these systems, region-of-interest segmentation is currently the rate-limiting step.^75^

We corroborate the findings of Vaassen et al.,^28^ who likewise reported the APL and surface DSC to be superior correlates with correction time. Importantly, our methodology differs from Vaassen et al. in that we used an autosegmentation algorithm that was not optimized to segment thoracic cavity volumes in CT scans from patients with NSCLC, whereas Vaassen et al. used a commercial atlas-based tool and a commercial prototype deep learning tool. The good correlation between the APL and surface DSC and correction time in our study suggests that these metrics may be robust even when evaluating autosegmentation tools that are not highly optimized for their tasks. In contrast, other metrics may degrade in this circumstance. For example, surface distance metrics performed dramatically worse in our study than in Vaassen et al. The maximum, 99^th^, and 98^th^ percentile HDs were worse correlates with correction time than the surface DSC even at an impractically high error tolerance (10 mm). Given the popularity of the HD as a measure of autosegmentation goodness, this alone is an informative result.

Autosegmentations have achieved unprecedented spatial similarity to reference segmentations^29,35,36,51,71^ and improved computational efficiency^37,43,47,76^ since deep learning’s^77^ emergence in 2012.^78^ Deep learning algorithms should be trained on data representing the spectrum of clinical variation, but the practical consequences of deploying algorithms that are not trained on diverse data remains an outstanding question. Our methodology permits an interesting case study in the time-savings value of deep learning autosegmentation tools that are deployed on classes of data that are underrepresented in the algorithms’ training data, since our autosegmentation algorithm was not trained on CTs from patients with NSCLC. We expected that autosegmentation spatial similarity losses due to unseen, cancer-induced anatomic variation would prolong the time required to correct autosegmentations. Rather, we made the interesting observation that clinical variation did not always cost time. Presumably, manual segmentation tools such as adaptable brush sizes and segmentation interpolation were enough to buffer similarity losses.

It is a limitation of this study that autosegmentation corrections were delineated by a fourth-year medical student, but all medical student segmentations underwent subsequent vetting by a radiation oncologist or radiologist and showed very high agreement with physician-corrected segmentations. Furthermore, we acknowledge that our conclusions are limited to the context of thoracic cavity segmentation and should be replicated for clinical autosegmentation tasks across medical domains.

## CONCLUSION

The APL, FNPL, and surface DSC captured the time-saving benefit of thoracic cavity autosegmentation better than traditional metrics. They correlated strongly with correction time even when controlling for clinical variables known to associate with correction time. This suggests that they may be preferred metrics for optimizing and evaluating autosegmentation algorithms that are intended to save clinicians time.

## Data Availability

Radiologist or radiation oncologist-vetted thoracic cavity segmentations are publicly available as the “Thoracic Volume and Pleural Effusion Segmentations in Diseased Lungs for Benchmarking Chest CT Processing Pipelines” analysis dataset through The Cancer Imaging Archive

https://doi.org/10.7937/tcia.2020.6c7y-gq39

## ACKNOWLEDGMENTS

The authors thank Dr. Femke Vaassen for correspondence clarifying the calculation of the added path length.

